# Safety, Feasibility, and Preliminary Clinical Findings of an Oral Polypore Fungi Combination with Mild-to-Moderate COVID-19: A Randomized, Placebo-Controlled Phase I Clinical Trial

**DOI:** 10.64898/2026.06.01.26354267

**Authors:** Gordon Saxe, Andrew Shubov, Christine N. Smith, Shahrokh Golshan, Tatyana Shekhtman, Stephen Wilson, Daniel Slater, Zolton J. Bair, Chase Beathard, Renee A. Davis, Lauray MacElhern, Lan K. Kao, Phoebe Senowitz, Natalie Gosnell, David Buchholz, Hector Aguilar-Carreno

## Abstract

**Background:** Use of fungal mycelia, which have innate immunity-enhancing and antiviral properties, constitutes a novel early-wave, low-cost, at-home strategy in the early critical months of an outbreak when treatments or mass medical countermeasures are not yet available and the public health recommendations advise quarantining at home.

**Methods:** We evaluated safety and feasibility of a fungal mycelial combination (Fomitopsis officinalis and Trametes versicolor, FoTv) for treatment of COVID-19 and explored its antiviral effects and potential to reduce symptoms. In a randomized, double-blind, placebo-controlled (26 FoTv, 24 Placebo), dual site (UCSD/UCLA medical centers) clinical trial conducted under an active FDA Investigational New Drug (IND) application, we examined non-hospitalized patients diagnosed with mild-to-moderate COVID-19 ≤ 96 hours, and experienced symptom onset ≤ nine days, before enrollment.

**Results:** FoTv was safe, well-tolerated, and feasible for COVID-19 treatment. Mean values for renal, hepatic, and coagulation markers for both the FoTv and Placebo groups remained within normal levels. Unlike the Placebo, the FoTv group exhibited a minor reduction in estimated glomerular filtration rate across the treatment period, but both groups were equally likely to transition from normal to abnormal levels across the treatment period. Treatment adherence was high (97% FoTv, 95% Placebo) and dropout was low (0% FoTv, 4% Placebo). Exploratory analyses indicated that FoTv significantly reduced the number and severity of symptoms, particularly sore throat/cough, as well as the COVID-19 biomarker, C-reactive protein. A complementary *in vitro* study indicated that FoTv reduced SARS-CoV-2 (pseudovirus) cellular infection.

**Conclusions:** FoTv was safe, feasible, reduced viral cellular infection, and may reduce symptoms of COVID-19 associated with viral transmission. In our recent companion clinical trial, examining FoTv as a COVID vaccine adjunct, we observed a significant reduction in short-term vaccine side effects while preserving—and potentially increasing—long-term anti-SARS-CoV-2 antibody levels over six months (Saxe et al., 2026, BMC Immunology). Taken together, these studies elucidate a blueprint for a novel two-phase public health pandemic intervention. This blueprint involves use of a botanical immunomodulatory product, during the early critical months of an outbreak and, once vaccines become available, to improve their uptake and effectiveness.

**Trial Registration:** Clinicaltrials.gov registration:NCT04667247.

## INTRODUCTION

### COVID-19: Lessons and guidance for future pandemics

The COVID-19 pandemic underscored the need for effective public health strategies to fill the therapeutic gap that may exist after widespread viral exposure but before mass medical countermeasures become available. Effective low-cost and accessible strategies, particularly if made available for individuals quarantining at home, could reduce morbidity, mortality, and global disruption. Although effective strategies now exist for managing SARS-CoV-2, emerging variants and other infectious viral diseases (e.g., bird flu, Zika virus, Hantavirus, dengue fever, etc.) continue to pose significant risks to global public health.

During the COVID-19 pandemic, vaccines and antiviral medications constituted the first lines of defense. However, this approach may not be fully reliable in the future because development of relevant vaccines and antivirals may not be able to keep pace with the emergence of new viral variants and diseases. In addition, vaccines are usually only effective if taken prior to disease exposure. Similarly, antivirals may be less effective unless they are taken during a limited time window (prior to or shortly after exposure or symptom onset) and may encounter drug resistance because of misuse or overuse.

Furthermore, vaccines and antiviral interventions may be limited by cost, supply constraints, or distribution challenges. Complementary approaches, particularly the use of accessible, shelf-stable, medical-grade botanical products, may help reduce these population-level risks especially because they may be more acceptable to and incur less resistance from the population.

To the extent that complementary approaches support innate baseline immune resiliency, they could provide a layer of defense not only against emerging variants of SARS-CoV-2, but also against other infectious viral diseases with global pandemic potential. Among these, the highly pathogenic avian influenza A, caused by a viral subtype of H5N1, is of growing concern. It has been detected in more than 500 avian and mammalian species worldwide^1,2^ with sporadic transmission to humans^3^ with symptoms in humans ranging from mild (e.g., conjunctivitis, flu-like symptoms) to severe or fatal outcomes (e.g., respiratory failure). Between January 2003 and April 2024, there were 896 reported cases across 24 countries, resulting in a case-fatality rate of approximately 52%^4^. The expanding host range and increasing evidence of cross-species transmission raise concerns about the potential for sustained human-to-human transmission.

### Host-directed innate immune enhancement and antiviral properties of polypore fungi, particularly Turkey tail and Agarikon

Fungi offer a novel, low-cost, scalable strategy to fill the therapeutic gap (between widespread exposure and medical countermeasures) because they both enhance innate immunity and are known to have a variety of antiviral properties. Unlike pathogen-specific antivirals, fungal immunomodulators act through conserved host pattern recognition receptors, particularly Dectin-1 and the Toll-like receptor (TLR) family, which recognize fungal cell wall components such as β-glucans and initiate NF-κB-mediated innate immune activation, promoting broad, nonspecific antiviral immunity ^5^. These host-directed effects may be complemented by the direct antiviral activities reported for polypore fungi.

Turkey tail *(Trametes versicolor,* Tv [syn. *Coriolus versicolor*]) is a widely studied polypore fungus with a long history of use in traditional medicine along with numerous clinical trials supporting the bioefficacy of its mycelium^6^. Interest in Tv has been growing because of its antiviral^7,8^ and immunomodulatory^9–11^ effects. Flow cytometry and cytokine profiling demonstrated that treatment with Tv mycelium resulted in notable changes in activation markers on human monocytes and lymphocytes and influenced the profile of pro-inflammatory, anti-inflammatory, and antiviral cytokines^10^. Tv has consistently exhibited wide-ranging antiviral activity in numerous studies, including potent anti-influenza effects^12–14^. In comparative analyses across multiple mushroom species, Tv mycelium emerged as the most promising candidate for both anti-influenza and anti-herpetic applications, likely due to its strong therapeutic efficacy and low toxicity ^15,16^. In addition, mechanistic studies of polysaccharide-K (PSK) suggest that fungal immunomodulation extends beyond β-glucans alone. While β-glucans engage Dectin-1, a distinct lipid fraction within PSK was identified as the TLR2 agonist, with these components acting synergistically to enhance innate immune activation^17^.

Moreover, PSK induces TNF-α and interleukin-6 (IL-6) secretion in murine macrophages through TLR4, with the response abolished in TLR4-deficient cells and blocked by TLR4 antibody pretreatment ^18^. More broadly, in vivo treatment of honeybees with Tv mycelium resulted in a substantial reduction in viral titers, including a 75-fold decrease in Lake Sinai virus, indicating cross-species antiviral activity in a real-world animal model^8^.

Agarikon (*Fomitopsis officinalis,* Fo [syn. *Laricifomes officinalis*]) is another polypore fungus with broad-spectrum antiviral, antibacterial, and immunomodulatory effects attributed to both mycelium and fruit bodies^19–21^, demonstrating its medicinal potential^22^. Mycelial extracts of Fo neutralized influenza A viruses, including H5N1, *in vitro* and significantly reduced infectious viral titers at non-cytotoxic concentrations^23^. *In vitro* activity of Fo has been shown against a range of pathogens, including viruses such as HSV, H5N1, H3N2, and Orthopoxviruses^23–26^, as well as bacteria such as *Staphylococcus aureus* and *Mycobacterium tuberculosis*^25,27^. Complementing its antimicrobial and antiviral properties, triterpenoids from Fo also showed anti-inflammatory effects in RAW 264.7 murine macrophages and have elicited NF-κB mediated immune-engaging effects *in vivo* in a zebrafish cancer model^28–30^.

Transcriptomic analysis of human peripheral blood mononuclear cells (PBMCs) treated with the same Fo mycelium strain used in the present clinical study found that Fo modulates expression of Dectin-1 (CLEC7A) as well as multiple TLR family members (TLR4, TLR5, TLR7) and their downstream regulators, with the direction of engagement shifting between resting and LPS-challenged conditions^31^. Recent genomic characterization of Fo has revealed numerous biosynthetic gene clusters which could potentially produce a diverse array of secondary metabolites underlying these reported bioactivities, including terpenes and polyketides^32^.

### Polypore fungi in the management of COVID-19

In the early stages of the COVID-19 pandemic, we explored ways to employ integrative medicine, particularly the use of fungi, as part of COVID-19 prevention and therapy. One strategy was to use mushroom mycelium as a vaccine adjunct to prevent the likelihood of contracting COVID-19^33^, an approach that had been previously explored in humans for influenza B^34^, and in the veterinary literature for other diseases^35–37^. Specifically, we recently carried out a randomized clinical trial^33^ of FoTv, a 50/50 blend of mycelia of Fo and Tv mycelia, harnessing the unique and potentially complementary benefits of each component. We demonstrated that it was a safe, well-tolerated, and feasible adjunct to COVID-19 vaccination. In previously unvaccinated individuals without previous COVID-19 infection, FoTv in conjunction with COVID-19 vaccination was associated with significantly reduced vaccine side effects while preserving, or potentially enhancing, Spike and receptor-binding domain (RBD) antibody titers over a six-month period^33^. Another strategy was to employ fungi to reduce the morbidity and mortality in individuals who had contracted COVID-19. Both approaches (prevention and therapy) work by enhancing and supporting the innate immune system and could help society achieve herd immunity more quickly and with less harm.

We investigated safety, feasibility of use, and preliminary effects of fungi on active disease, responding to the urgent real-world need at the beginning of a pandemic to find or develop effective treatments for infected individuals. The primary objective was to examine, in patients with mild-moderate COVID-19 self-quarantining at home, whether oral treatment with FoTv was safe and feasible. We hypothesized that treatment with FoTv, versus Placebo, would be safe and feasible. We also explored potential efficacy of FoTv in reducing COVID-related symptoms and biomarkers, and SARS-CoV-2 viral load. A secondary objective was to characterize the effects of FoTv on SARS-CoV-2 viral cellular infection. We therefore further hypothesized FoTv would significantly inhibit *in vitro* viral entry into Vero-TMPRSS2 (mammalian epithelial) cells versus control condition.

## METHODS

### Study Design

This was a 14-day, double-blind, placebo-controlled, dual-site randomized clinical trial. Food and Drug Administration (FDA) approval was obtained prior to recruitment and enrollment. Participants were recruited from University of California San Diego (UCSD) or University of California Los Angeles medical centers and their local communities and provided written consent at UCSD. The trial protocol can be accessed at ClinicalTrials.gov, Identifier: NCT04667247^38^ and this study adheres to CONSORT guidelines (see CONSORT Checklist).

### Participants

A planned recruitment of 66 (53 completers) was estimated based on the number needed to assess safety and feasibility, which included a 20% dropout rate. Sex was self-reported in response to an open-ended query.

### Eligibility Criteria

Inclusion: (1) Diagnosed with COVID-19 within the prior 96 hours; (2) onset of symptoms within a maximum of nine days prior to enrollment; (3) 18 years or older.

Exclusion: (1) Known renal or liver failure; (2) acute symptoms that required hospitalization (3) current use of investigational agents to prevent or treat COVID-19; (4) prisoners; or (5) pregnant or breastfeeding women.

Participants were asked to minimize alcohol and cannabis use during the study period. Use of monoclonal antibodies, supplements, hydroxychloroquine, ivermectin, and vaccines were permitted throughout the study.

### Randomization and Masking

After enrollment (Day 0), participants were randomly assigned in a 1:1 ratio to receive FoTv or Placebo starting on Day 1 (Figure 1). Prior to recruitment, the statistician (S.G.) created a randomization table using block sizes of 4 and 6 (SPSS software) and then randomization status was directly communicated to the pharmacist, who shipped the capsules to participants without direct communication. Only the statistician and pharmacist were unblinded to participant randomization. FoTv and Placebo capsules were visually identical and distributed in unlabeled containers.

**Figure 1.**
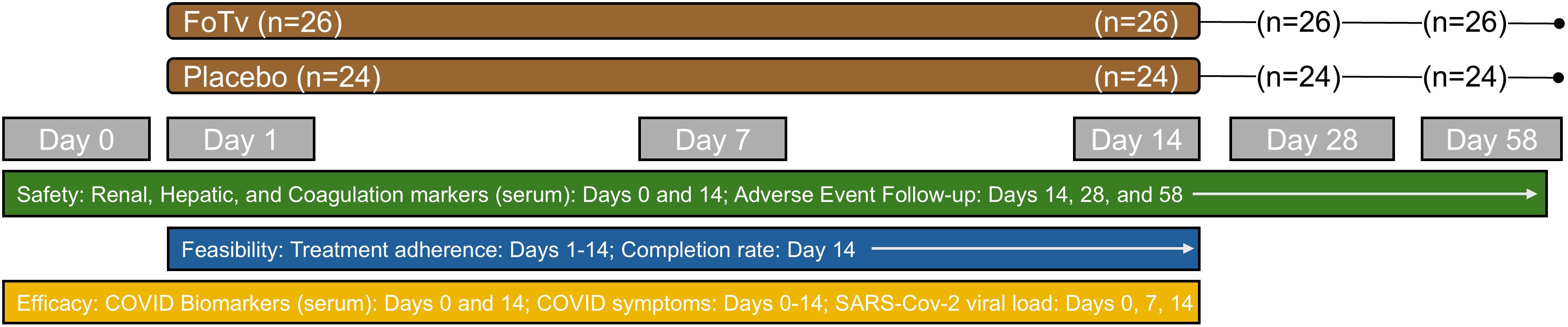
Design of Clinical Trial. Participants with COVID-19 were randomized to receive either a 14-day regimen of FoTv or Placebo on Day 0 (baseline). Renal, hepatic, and coagulation markers from serum were examined for safety on Days 0 and 14 and adverse events were assessed at follow-up visits on Days 14, 28, and 58. Feasibility analyses examined treatment adherence (Days 1-14) and completion rate at Day 14. Preliminary efficacy analyses examined COVID biomarkers (serum: Days 0 and 14), COVID symptoms (self-report: Days 0-14), and SARS-CoV-2 viral load (nasal swab: Days 0, 7, and 14).

### Materials

FoTv consisted of dried FoTv mycelium and the fermented brown rice substrate on which it was cultivated, and inert Placebo of dried cooked organic brown rice. Fo and Tv mycelia were independently cultivated through solid state fermentation on an organic brown rice substrate. Each myceliated fermented substrate ingredient was frozen, dried, and milled into a powder before being combined evenly and encapsulated in 500-mg pullulan capsules to produce the FoTv formulation. FoTv and Placebo were developed, encapsulated to a cGMP standard, and provided by Fungi Perfecti, LLC, which was granted Investigational New Drug (IND) status and approved for study in a Phase I clinical trial by the FDA. As required by the FDA, certificates of analysis were obtained for Fo, Tv, and Placebo. Capsules were packaged and appeared identical to untrained study participants. FoTv/Placebo dosage was eight capsules three times daily (TID) for 14 consecutive days. The daily dosage was based on a prior study of effects of Tv on immune response^11^ and was identical to that employed in a clinical trial of individuals who received FoTv for four days as an adjunct to COVID-19 vaccination.^33^ A treatment duration of 14 days was selected to minimize subject burden and maximize adherence, while timing delivery to the typical 1-2 week duration of COVID-19 symptoms at the time of study design^39^.

### Procedure

Participants in both groups received mobile phlebotomy services to collect blood samples on Day 0 and Day 14, provided mid-turbinate nasal swabs on Days 0, 7 and 14 (to assess viral load), and completed a daily symptom diary from Day 0-14 (Figure 1). Following the 14-day treatment period, follow-up phone calls were conducted on Days 28 and 58 to review symptoms and identify any emergency department or hospital visits. Compliance with study medication was monitored by a combination of daily diary, routine phone calls by the study team, and photo evidence of empty bottles at study completion.

### Viral entry assay using SARS-CoV-2 spike-pseudotyped VSV

Viral entry was measured using a modified vesicular stomatitis virus (VSV) that carried the SARS-CoV-2 spike (S) protein on its surface. This type of virus, a “pseudovirus”, is useful for studying viral entry because it can enter cells but is not able to complete a full round of virus production on its own^40–42^. The virus also contained a *Renilla* luciferase (Rluc) reporter gene, which generated a light signal after the virus entered cells. This allowed viral entry to be measured by luminescence. Because the normal VSV glycoprotein gene was removed, the virus could undergo only a single round of infection and could not produce new infectious virus unless a glycoprotein (SARS-CoV-2 Spike) was externally sourced to create the pseudovirus.

For assay optimization, a range of pseudovirus input doses was tested to identify conditions that produced a reporter signal within the linear range of detection. For inhibition experiments, the selected amount of pseudovirus was incubated with FoTv at a final concentration of 0.01 mg/mL for 1 hour before infection. Because FoTv was dissolved in dimethyl sulfoxide (DMSO), an equivalent volume of DMSO alone was included as a vehicle control. As a validation control, we also included an uninfected control condition in which cells were incubated with plain media. The resulting pseudovirus mixtures were added to Vero-TMPRSS2 (mammalian epithelial) cells and incubated for 24 hours under standard culture conditions. After incubation, the cells were lysed and Rluc activity was measured according to the manufacturer’s protocol (Promega Rluc Assay System). Luminescence was then read on a Tecan plate reader as an indicator of pseudovirus entry. Three independent replicates of each condition were performed.

### Outcome Variables

#### Primary Objective: Randomized Clinical Trial

##### Safety

(1) Adverse event (hospitalizations; ER or urgent care visits) frequency across Days 1-14; and (2) markers of serum renal and hepatic function and of coagulation on Days 0 and 14. Participants were classified as having normal vs. abnormal renal function using markers including adjusted estimated glomerular filtration (eGFR)^43^, as well as sodium, chloride, and blood urea nitrogen (BUN). Classification of normal vs. abnormal hepatic function was based on serum total protein, albumin, alkaline phosphatase (ALP), Aspartate Aminotransferase (AST), Alanine Transaminase (ALT), and total bilirubin. Classification of normal vs. abnormal coagulation was based on prothrombin time, activated partial thromboplastin time (APTT), and international normalized ratio. Cutoff scores for normal vs. abnormal classifications appear in Supplemental Table 1.

##### Feasibility

(1) Completion rate at Day 14 (end of safety assessment period); and (2) Treatment Adherence= (capsules taken/assigned) * 100%.

Exploratory Efficacy: (1) number, severity, and duration of COVID-19 symptoms as assessed by daily questionnaires, (2) serum COVID-19 biomarkers associated with COVID-19 severity [high-sensitivity Troponin-T Gen 5 (hs-cTnT), Ferritin, C-reactive protein (CRP), D-dimer, Erythrocyte sedimentation rate (ESR), and Lactate dehydrogenase (LDH)], and (3) SARS-CoV-2 viral load (polymerase chain reaction [PCR] cycle threshold value--a measure of detectable viral genetic material).

Participants were asked to rate their symptoms on a scale of 0 (none or absent) to 4 (very severe) across 12 symptoms commonly associated with COVID-19: fever, fatigue, muscle aches, shortness of breath, shortness of breath upon exertion, cough, sore throat, stuffy nose, runny nose, loss of taste, loss of smell, and headache. Symptom summary outcome measures were also computed for each day by (1) counting the total number of symptoms reported (Symptom Count), and (2) summing the severity ratings of all symptoms reported (Symptom Severity). Symptom Duration was computed by identifying the study day (Days 1-14) on which Symptom Count (or Severity) had decreased by 50% compared to Day 0. Results for individual symptoms and symptom summary outcome measures are reported in Table 3 and Supplemental Table 2.

#### Secondary Objective: *In Vitro* Experiment

Vero-TMPRSS2 cells were lysed, and *Renilla* luciferase activity was measured according to the manufacturer’s instructions as a readout of viral entry. Luminescence values were used to quantify relative pseudovirus entry into cells.

#### Data Management and Analysis

Data were managed and statistical analyses performed by Krupp Center for Integrative Research (KCIR) biostatistics and data management core, using the KCIR Data System. Each participant was assigned a sequential participant number. Baseline group demographic and clinical feature comparability was tested using Analysis of Variance and Chi-square analyses. Descriptive analyses assessed data distribution, normality, and homogeneity. Missing data and group dropout rate were examined for randomness. Data were analyzed in SPSS (v.28) using two-tailed statistical tests (differences considered statistically significant for p values <0.05).

##### Feasibility

Feasibility was determined by obtaining a minimum of 80% completion/adherence rates.

##### Safety

Linear mixed-effects (LME) analyses compared serum levels between one grouping factor (Treatment Group: FoTv vs. Placebo) across two timepoints (Days: 0 and 14). Differences between groups across days were examined using in terms of the percentage (±95% CI) of participants transitioning from a normal serum level of each marker on Day 0 to an abnormal level on Day 14.

##### Exploratory Efficacy

We computed means, standard deviations, effect sizes (partial eta squared, η^2^_p_), and power analyses for outcome variables. Preliminary efficacy analyses used linear mixed-effects (LME) analyses. The first set of analyses examined symptoms. We compared symptoms (Count or Severity) between one grouping factor (Treatment Group: FoTv vs. Placebo) across 15 timepoints (Days: 0-14). Symptom Duration was tested using a between-subjects Analysis of Variance (ANOVA) examining Treatment Group. We also examined whether symptom outcome measures were related to vaccination status (vaccinated vs. unvaccinated). The second set of analyses compared serum levels of COVID-19 biomarkers between one grouping factor Treatment Group (FoTv vs. Placebo) across two timepoints (Days: 0 and 14). The third analysis compared SARS CoV-2 Viral Load between one grouping factor Treatment Group (FoTv vs. Placebo) across three timepoints (Days: 0, 7, and 14). For each analysis, the main effects of Group, Day, and the Group by Day interaction were examined.

The standards for serum levels for five serum outcomes (total protein, sodium, BUN, ferritin, and D-dimer) were different across the two recruitment sites. For these outcomes, data were z-transformed prior to mixed-effects analyses.

*In vitro* luminescence data were analyzed using Analysis of Variance comparing FoTv to DMSO on three independent biological samples. Data from the uninfected condition are also shown.

## RESULTS

### Sample Characteristics

Participants were enrolled between December 2020 and March 2022, with the last follow-up visit in August 2022. Fifty participants were enrolled, 26 receiving FoTv and 24 Placebo (Figure 1, Figure 2). Mean participant age was 45.4 ± 14.4 years. Participants were: 64% female, 84% Caucasian, 8% Asian, 2% African American, 2% Native American, and 4% other race; 27% reported Hispanic ethnicity.

**Figure 2.**
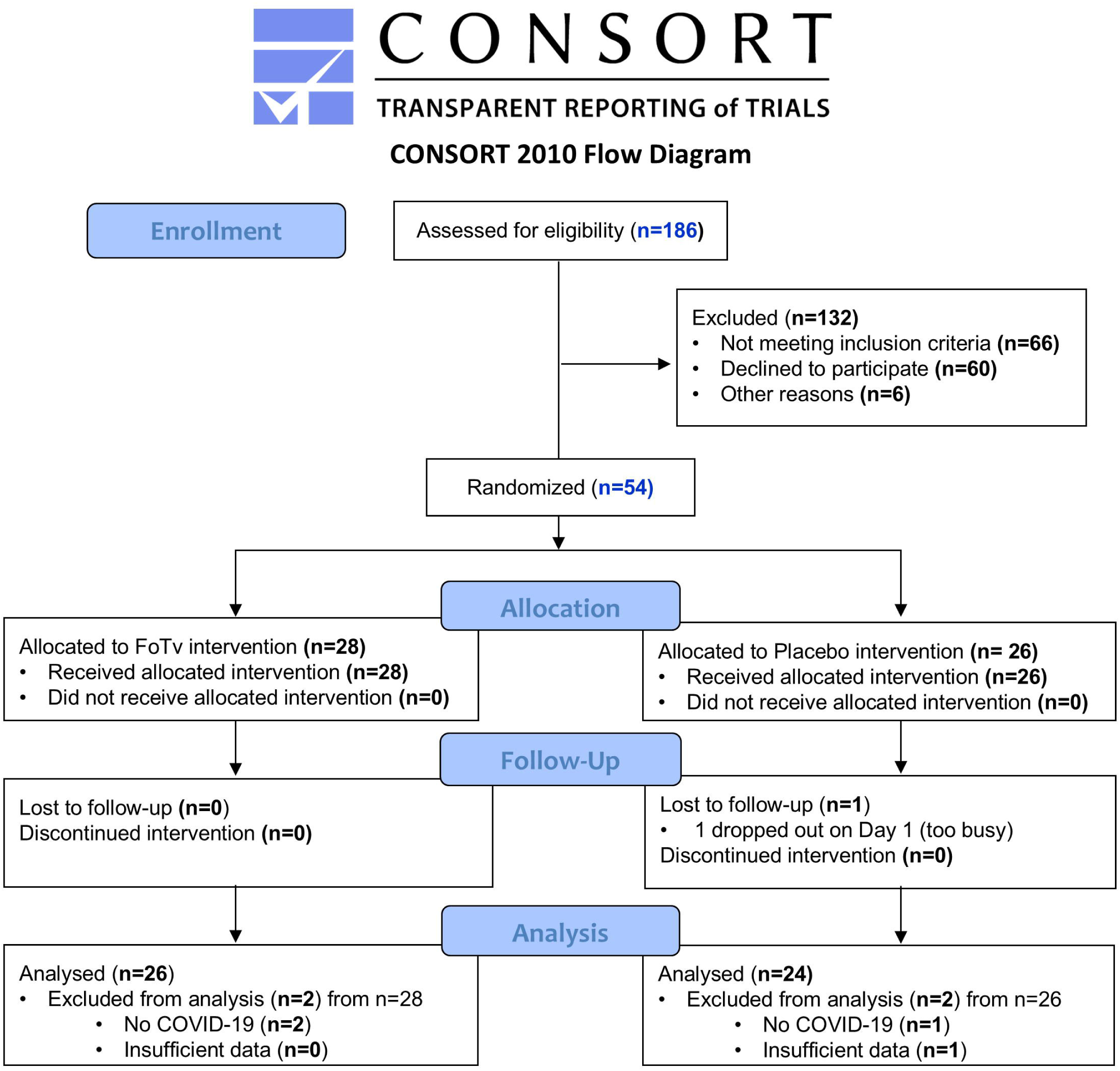
CONSORT Diagram.

At baseline (Day 0), there were no significant differences between FoTv and Placebo groups in demographic or clinical characteristics, presence of cardiovascular disease, hypertension, or diabetes, or the number of days between symptom onset and start of treatment (Day 1), indicating effectiveness of the randomization procedure (Table 1).

**Table 1.** Patient Demographics and Baseline Clinical Characteristics.

| Characteristic | FoTv | Placebo | Statistic | df | p value |
| --- | --- | --- | --- | --- | --- |
|  | Mean/Count (SD/%) |  |  |  |  |
| Number of Participants | 26 | 24 |  |  |  |
| Age | 43.3 (11.1) | 47.7 (17.3) | 1.188 | 1,48 | 0.281 |
| Body Mass Index | 25.8 (5.1) | 25.7 (5.3) | 0.002 | 1,45 | 0.964 |
| Sex (female) | 16 (61.5%) | 16 (66.7%) | 0.142 | 1 | 0.706 |
| Race |  |  | 3.020 | 4 | 0.554 |
| Asian | 3 (11.5%) | 1 (4.2%) |  |  |  |
| Caucasian | 20 (76.9%) | 22 (91.7%) |  |  |  |
| African | 1 (3.8%) | 0 (0.0%) |  |  |  |
| Native American | 1 (3.8%) | 0 (0.0%) |  |  |  |
| Other | 1 (3.8%) | 1 (4.2%) |  |  |  |
| Ethnicity |  |  |  |  |  |
| Hispanic | 7 (28.0%) | 6 (25.0%) | 0.057 | 1 | 0.812 |
| Past Medical History |  |  |  |  |  |
| Cardiovascular Illness | 1 (4.0%) | 1 (5.0%) | 0.026 | 1 | 1.000 |
| Diabetes Mellitus | 0 (0%) | 3 (15.0%) | 4.018 | 1 | 0.080 |
| Hypertension | 1 (4.0%) | 4 (20.0%) | 2.880 | 1 | 0.155* |
| COVID-19 status |  |  |  |  |  |
| Vaccinated | 23 (88.5%) | 17 (70.8%) | 2.424 | 1 | 0.164* |
| Use of MAT | 2 (7.7%) | 1 (4.2%) | 0.297 | 1 | 0.862* |
| Days to enrollment | 5.5 (1.7) | 5.8 (1.8) | 0.428 | 1,48 | 0.516 |

*Note.* Statistic refers to $\chi^2$ for categorical variables and F value (ANOVA) for continuous variables. \*Due to the small sample size for cells in these analyses, probability values for Fischer's exact tests are reported in lieu of chi-squared tests. MAT, monoclonal antibody treatment.

The means of the two groups were not significantly different at baseline with regard to any laboratory data, except for BUN (F_(1,49)_ = 4.222, p = 0.045), which was significantly higher in the Placebo than the FoTv group (Table 2).

**Table 2.** Serum Safety Findings at Day 0 (baseline) and at Day 14.

|  | Comparison of Laboratory Data across Groups and Days |  |  |  |  | Percentage Transitioning from Normal to Abnormal Function from Day 0 to Day 14 |  |  |  |
| --- | --- | --- | --- | --- | --- | --- | --- | --- | --- |
|  | Day 0 |  | Day 14 |  | Group by Day Effect<br>p value |  |  |  |  |
|  | FoTv | Placebo | FoTv | Placebo |  | FoTv |  | Placebo |  |
|  | Mean (SD) | Mean (SD) | Mean (SD) | Mean (SD) |  | % | 95% CI | % | 95% CI |
| *Hepatic Function* |  |  |  |  |  |  |  |  |  |
| Total Protein<sup>1</sup> | 0.24 (0.90) | -0.03 (1.07) | 0.06 (0.82) | -0.29 (0.87) | 0.417 | 0 | 0.0-15.4 | 0 | 0.0-14.8 |
| Albumin | 4.57 (0.36) | 4.52 (0.27) | 4.64 (0.29) | 4.56 (0.29) | 0.690 | 4.0 | 0.1-20.4 | 0 | 0.0-14.8 |
| Alkaline Phosphatase | 75.64 (38.58) | 76.29 (22.89) | 68.50 (20.09) | 77.54 (19.80) | 0.109 | 0 | 0.0-14.2 | 0 | 0.0-14.8 |
| AST | 23.50 (7.81) | 22.92 (6.41) | 20.73 (4.94) | 20.54 (4.40) | 0.841 | 0 | 0.0-13.7 | 0 | 0.0-14.2 |
| ALT | 28.15 (21.75) | 22.92 (10.90) | 22.62 (11.88) | 19.79 (7.16) | 0.612 | 4.3 | 0.1-21.9 | 0 | 0.0-14.8 |
| Total Bilirubin | 0.42 (0.19) | 0.37 (0.13) | 0.55 (0.24) | 0.42 (0.19) | 0.174 | 0 | N/A | 0 | N/A |
| *Renal Function* |  |  |  |  |  |  |  |  |  |
| Adjusted eGFR | 111.94 (20.47) | 101.73 (22.89) | 104.28 (20.42) | 103.87 (21.26) | 0.010 | 28.6 | 11.3-52.2 | 0 | 0.0-23.2 |
| Sodium<sup>1</sup> | -0.11 (1.19) | -0.06 (1.02) | -0.26 (0.91) | 0.46 (0.68) | 0.063 | 4.3 | 0.1-21.9 | 0 | 0.0-14.2 |
| Chloride | 101.77 (2.96) | 101.63 (2.57) | 102.35 (1.77) | 102.63 (1.69) | 0.625 | 4.3 | 0.1-21.9 | 0 | 0.0-14.8 |
| BUN<sup>1</sup> | -0.46 (0.90) | 0.11 (1.06) | -0.06 (0.72) | 0.46 (1.10) | 0.607 | 0 | 0.0-15.4 | 0 | 0.0-15.4 |
| *Coagulation* |  |  |  |  |  |  |  |  |  |
| Prothrombin Time | 11.52 (0.77) | 11.45 (0.93) | 11.34 (0.57) | 11.34 (1.15) | 0.846 | 0 | 0.0-14.8 | 14.3 | 3.0-36.3 |
| APTT | 32.38 (3.10) | 32.23 (2.98) | 32.11 (2.87) | 31.51 (3.08) | 0.560 | 4.5 | 0.1-22.8 | 5.0 | 0.1-24.9 |
| INR | 1.07 (0.06) | 1.05 (0.06) | 1.05 (0.06) | 1.03 (0.06) | 0.707 | -- | -- | -- | -- |

Note. Linear Mixed Effects models (2X2) were used to test the Group by Day interactions. For safety, the Normal to Abnormal transition analysis reflects the percentage of participants with normal function at Day 0 that transitioned to abnormal at Day 14 compared to participants with normal function at both Days. <sup>1</sup>Recruitment site laboratories used different standards, therefore z scores were used. -- normal range not provided by laboratories. N/A = the 95% confidence intervals could not be computed for Total Bilirubin because all participants were normal at both Day 0 and Day 14. AST, Aspartate Aminotransferase; ALT, Alanine Transaminase; APTT, activated partial thromboplastin time; BUN, Blood Urea Nitrogen; CI, Confidence Interval; eGFR, estimated glomerular filtration rate; INR, International Normalized Ratio.

Nevertheless, some participants had abnormal baseline markers of hepatic (Total Protein [four FoTv participants, one Placebo participant]; Albumin [one FoTv, two Placebo], ALP [three FoTv and one Placebo], AST [two FoTv, zero Placebo], and ALT [one FoTv, three Placebo]) or renal function (eGFR [two FoTv, 11 Placebo], Sodium [three FoTv, zero Placebo], Chloride [three FoTv, one Placebo], BUN [four FoTv, two Placebo]). Additionally, some participants had abnormal markers for coagulation (Prothrombin Time [two FoTv and two Placebo] and APTT [four FoTv and four Placebo]).

At baseline, mean viral load was 15.3 cycle threshold in the FoTv group and 14.9 cycle threshold in the Placebo group, demonstrating a high viral load in the population, as well as matched viral load across groups (F_(1,41)_ = 0.056, p = 0.814) (Table 3).

**Table 3.** Exploratory Efficacy Findings for COVID-19 Symptoms and Biomarkers, and SARS-CoV-2 Viral Load, at Day 0 (baseline) and at Day 14.

| | Comparison of Groups across Day 0 and Day 14 | | | | Group by Day Effect<br>p value | Effect Size<br>$\eta^2_p$ | Power |
| --- | --- | --- | --- | --- | --- | --- | --- |
|  | Day 0 |  | Day 14 |  |  |  |  |
|  | FoTv | Placebo | FoTv | Placebo |  |  |  |
|  | Mean (SD) | Mean (SD) | Mean (SD) | Mean (SD) |  |  |  |
| **COVID-19 Symptoms** |  |  |  |  |  |  |  |
| Symptom Count | 7.23 (2.30) | 6.88 (1.99) | 1.11 (1.66) | 2.17 (2.24) | 0.005 | 0.050 | 0.336 |
| Symptom Severity | 13.65 (6.62) | 11.08 (4.26) | 1.61 (2.78) | 2.92 (3.54) | <0.001 | 0.137 | 0.763 |
| **Individual COVID Symptoms** |  |  |  |  |  |  |  |
| Sore throat | 1.57 (1.20) | 0.59 (0.67) | 0.00 (0.00) | 0.00 (0.00) | <0.001 | 0.160 | 0.835 |
| Cough | 1.77 (0.99) | 1.37 (0.76) | 0.23 (0.51) | 0.42 (0.65) | <0.001 | 0.099 | 0.604 |
| Muscle aches | 1.39 (0.99) | 0.86 (0.90) | 0.13 (0.34) | 0.40 (1.00) | 0.001 | 0.022 | 0.171 |
| Headache | 1.09 (1.00) | 1.05 (0.84) | 0.00 (0.00) | 0.35 (0.67) | 0.003 | 0.040 | 0.281 |
| Runny nose | 1.35 (1.03) | 1.09 (0.97) | 0.09 (0.29) | 0.15 (0.37) | 0.022 | 0.132 | 0.746 |
| Stuffy nose | 1.87 (1.10) | 1.36 (1.14) | 0.22 (0.42) | 0.20 (0.41) | 0.038 | 0.032 | 0.233 |
| Fatigue | 1.70 (0.97) | 1.14 (0.80) | 0.30 (0.77) | 0.50 (0.95) | 0.084 | 0.028 | 0.204 |
| Loss of taste | 0.74 (1.29) | 0.73 (1.20) | 0.17 (0.49) | 0.45 (0.90) | 0.155 | 0.098 | 0.599 |
| Fever | 0.73 (0.96) | 0.58 (0.83) | 0.00 (0.00) | 0.00 (0.00) | 0.168 | 0.060 | 0.396 |
| Loss of smell | 0.70 (1.26) | 0.73 (1.20) | 0.48 (1.12) | 0.25 (0.72) | 0.189 | 0.025 | 0.190 |
| Shortness of breath upon exertion | 0.70 (1.06) | 0.73 (0.94) | 0.13 (0.46) | 0.20 (0.41) | 0.591 | 0.018 | 0.129 |
| Shortness of breath | 0.30 (0.64) | 0.23 (0.43) | 0.00 (0.00) | 0.15 (0.49) | 0.919 | 0.072 | 0.463 |
| **COVID-19 Biomarkers** |  |  |  |  |  |  |  |
| Troponin-T<sup>1</sup> | 5.88 (1.92) | 8.13 (6.11) | 7.32 (3.77) | 7.48 (5.77) | 0.029 | 0.100 | 0.598 |
| Ferritin<sup>2</sup> | 0.09 (0.99) | 0.05 (0.98) | 0.08 (1.18) | -0.23 (0.80) | 0.035 | 0.079 | 0.500 |
| C-reactive protein | 0.94 (1.16) | 0.55 (0.38) | 0.33 (0.48) | 0.37 (0.44) | 0.044 | 0.081 | 0.513 |
| D-dimer<sup>2</sup> | 0.24 (1.78) | 0.03 (0.46) | -0.20 (0.20) | -0.10 (0.52) | 0.283 | 0.017 | 0.136 |
| ESR | 12.54 (9.29) | 14.89 (13.78) | 11.13 (13.77) | 12.68 (10.41) | 0.765 | 0.004 | 0.069 |
| LDH | 186.08 (29.02) | 195.38 (41.48) | 195.77 (33.45) | 207.70 (37.92) | 0.884 | 0.000 | 0.052 |
| **SARS-CoV-2 Viral Load<sup>3</sup>** | **15.29 (4.93)** | **14.94 (4.66)** | **37.72 (5.15)** | **37.41 (4.78)** | **0.080** | **0.101** | **0.543** |

Note. Linear Mixed Effects models were used to test the Group by Day interactions for COVID-19 biomarkers (FoTv/Placebo X Days 0,14) and Symptoms (FoTv/Placebo X Days 0-14). Data for individual COVID-19 symptoms reflect severity ratings on a scale of 0 (none or absent) to 4 (very severe). <sup>1</sup>Data available only from UCSD site. <sup>2</sup>Recruitment site laboratories used different standards, therefore z scores were used. <sup>3</sup>Viral load is reported as cycle threshold values, which are inversely proportional to viral load. Full statistical reporting for COVID-19 symptoms and biomarkers appears in Supplemental Table 2. ESR, Erythrocyte Sedimentation Rate; LDH, lactate dehydrogenase. Effect sizes reflect partial eta squared ( $\eta^2_p$ ) obtained from repeated measures ANOVA (small effect $\approx 0.01$ , medium effect $\approx 0.06$ , large effect $\approx 0.14$ ) and the value reflects the percent of variance explained (e.g., 0.14 = 14% of variance explained).

### Feasibility

Feasibility analyses examined 186 participants for eligibility, of whom 132 were excluded and 54 enrolled with 28 originally randomized to FoTv and 26 to Placebo (Figure 2). In the Placebo group, one participant was lost to follow-up (on Day 1). In the FoTv group, none were lost to follow-up. Analysis of SARS-CoV-2 viral levels indicated that three participants did not have, or had already recovered from, COVID-19 upon enrollment (2 FoTv; 1 Placebo), so these participants were excluded from analysis. From the 51 (26 FoTv, 25 Placebo) remaining participants, 98.8% completed Days 0-14 (26 FoTv [100%], 24 Placebo [96%]). All participants who completed Day 14 also completed the Day 58 follow-up (Figure 1). Treatment Adherence was high (96.1% ± 5.1%, CI: 94.7-97.6%); and similar for FoTv (96.9% ± 4.7%, CI: 95.0-98.8%) and Placebo (95.3% ± 5.6%, CI: 93.0-97.7%).

### Safety

#### Sentinel Study (Required by FDA)

The FDA required close monitoring of the first six participants (3 FoTv, 3 Placebo) due to concern that polypore fungi (known to modify immune response) might inadvertently trigger cytokine storm, a sudden, inexplicable, severe, and potentially life-threatening immune response to infection that had been observed early in the course of the COVID-19 pandemic^44^. There was no indication of cytokine storm in these participants, and the FDA cleared the larger study to proceed.

#### Adverse Events

No hospitalizations, ER, or urgent care visits were reported in either group during the 14-day study period or across the 28- and 58-day follow-ups.

Out of the four participants with hypertension in the Placebo group, one reported abnormal blood pressure on Days 7 and 12. The one individual with hypertension in the FoTv group reported abnormal hypertension on Days 2 to 6 and 8 to 14. Four participants had hyperglycemia; the single participant in the FoTv group experienced no deviations outside of the normal range, whereas one of the three participants in the Placebo group had abnormal glucose at the baseline visit.

#### Renal Function, Hepatic Function, and Coagulation

There were no statistically significant differences between FoTv and Placebo groups from Day 0 to Day 14 for measures of hepatic or renal function or coagulation, except for adjusted eGFR (F_(1,45)_ = 7.186, p = 0.010), which exhibited a significant Group by Day interaction (Table 2). Follow-up tests revealed that eGFR significantly decreased across days in the FoTv group (F_(1,22)_ = 5.327, p = 0.031), but not in the Placebo group (F_(1,23)_ = 1.697, p = 0.206). Notably, there were no group differences in these eGFR levels at Day 0 or Day 14 (p values > 0.114).

We also examined whether the groups differed in the proportion of participants that transitioned from normal to abnormal from Day 0 to Day 14 (versus those that remained normal on both days). Importantly, there were overlapping CIs for the percentage of participants in each group transitioning from normal to abnormal function across this timeframe for both hepatic and renal function, as well as for coagulation (Table 2).

### Exploratory Efficacy

#### COVID-19 Symptom Scores

There were significant Group by Day (Days 0-14) interactions for Symptom Severity and Symptom Count (Table 3, Supplemental Table 2). In both cases, lower symptom severities (η^2^_p_= 0.137, power=0.763; F =16.560, p < 0.001), and counts (η^2^_p_= 0.050, power=0.336; F_(1,697)_=7.835, p = 0.005), were observed in the FoTv relative to the Placebo group (see Figure 3A for Symptom Count). Among the 12 COVID-19 symptoms, the severity of the following symptoms exhibited reductions evidenced by significant Group by Day interactions: sore throat (η^2^_p_= 0.160, power=0.835), cough (η^2^_p_= 0.099, power=0.604), muscle aches (η^2^_p_= 0.022, power=0.171), headache (η^2^_p_= 0.040, power=0.281), stuffy nose (η^2^_p_= 0.032, power=0.233), and runny nose (η^2^_p_= 0.132, power=0.746; Table 3 and Supplemental Table 2). Among these, there were particularly strong effects were observed for sore throat, runny nose, and cough.

**Figure 3.**
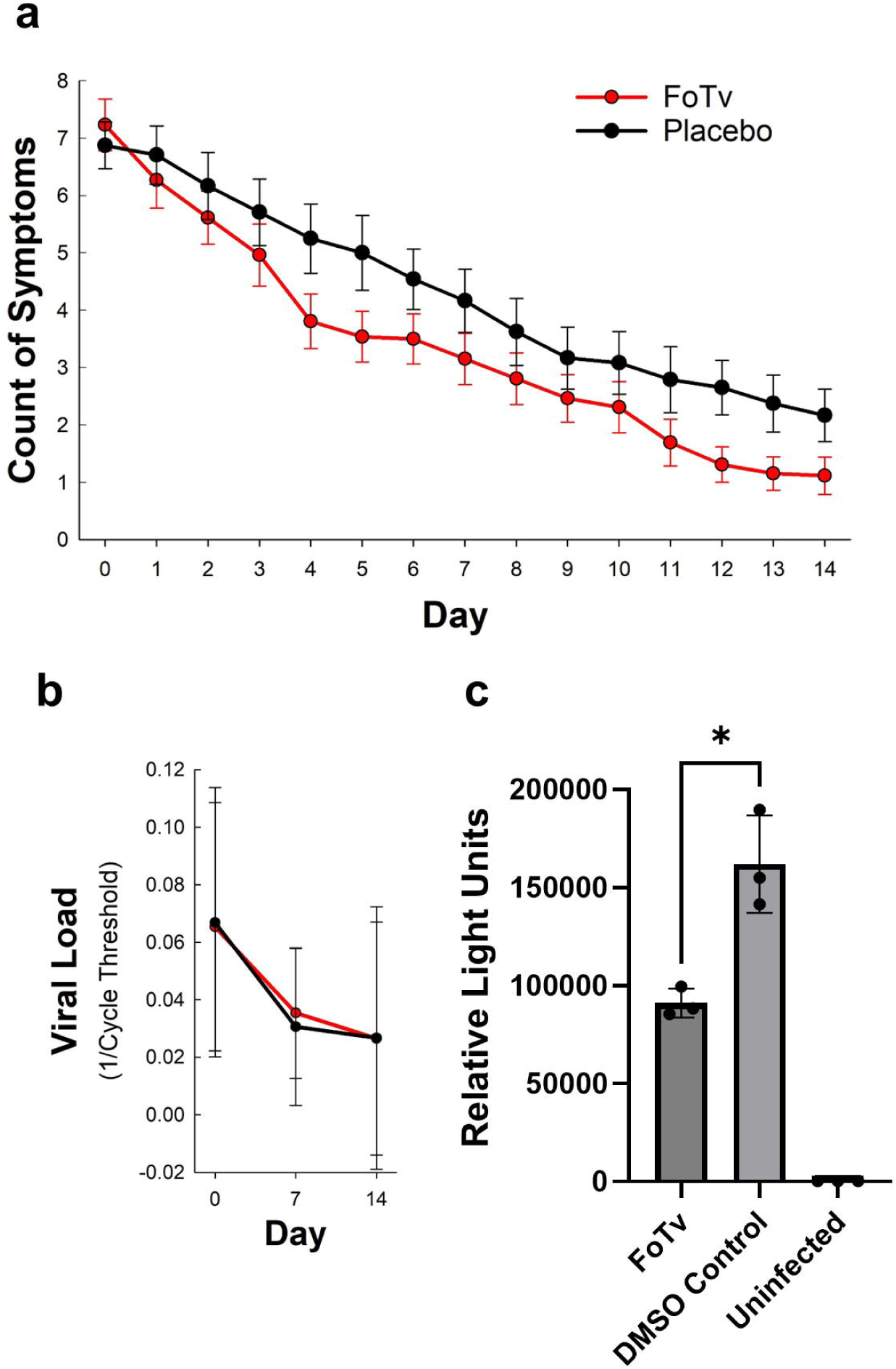
Efficacy Results for Symptoms, Viral Load in Humans, and *In Vitro* Viral Infection. **a.** Mean Symptom Count for the 12 COVID-19 symptoms assessed for FoTv (red, n=26) and Placebo (black, n=24) groups from Day 0 through Day 14. The FoTv group exhibited significantly fewer symptoms than the Placebo group across days (F-test, p = 0.005). Error bars show SEM. **b.** Mean nasal swab viral load at days 0, 7, and 14 via PCR. Viral load (i.e., detectable SARS-CoV-2 genetic material) significantly decreased across days (F-test, p < 0.001) and did so similarly for both groups (F-test interaction, p = 0.080). Error bars show SEM. **c.** Mean viral infection of TMPRSS2 cells with FoTv or control (DMSO) after 1-hour of incubation showed significant inhibition (F-test, *p = 0.009) of pseudotyped SARS-CoV-2 entry into TMPRSS2 cells (black circles show the three independent biological samples for each condition). Relative Light Units are non-standardized units that measure Renilla luciferase luminescence values as a quantification of relative pseudovirus entry into cells. Error bars show SD.

Exploratory analyses examining Vaccination Status (vaccinated/unvaccinated) by Group (FoTv/Placebo) by Day (Days 0-14) interactions revealed a strong trend for both Symptom Count (η^2^_p_= 0.003, power=0.063; F = 2.917, p = 0.055) and Symptom Severity (η^2^_p_= 0.004, power=0.072; F = 2.829, p = 0.060)(Supplemental Figure 1), though the effect sizes were small. These statistical findings should be interpreted with caution because of the small sample size of the unvaccinated group (5 FoTv, 7 Placebo). When vaccinated and unvaccinated subgroups were examined separately, the Group by Day interaction effect sizes were comparable for these subgroups and possibly larger in the unvaccinated group for Symptom Count (vaccinated: η^2^_p_= 0.041, power=0.221; unvaccinated: η^2^_p_= 0.090, power=0.147) and for Symptom Severity (vaccinated: η^2^_p_= 0.129, power=0.599; unvaccinated η^2^_p_= 0.131, power=0.199), For Symptom Count, the vaccinated FoTv group reached ≥50% reduction 1.3 days earlier than Placebo (FoTv: 5.7 days, Placebo: 6.9 days; η^2^_p_= 0.021, power=0.134), whereas the unvaccinated FoTv group reached ≥50% reduction in the number symptoms 5.1 days earlier than Placebo (FoTv: 5.8 days, Placebo: 10.9 days; η^2^_p_= 0.255, power=0.388). The findings were similar for Symptom Severity where vaccinated FoTv exhibited a 50% reduction in severity 1.2 days earlier than Placebo (FoTv: 5.1 days, Placebo: 6.3 days; η^2^_p_= 0.015, power=0.111) and unvaccinated FoTv exhibited a reduction 3.1 days earlier than Placebo (FoTv: 7.0 days, Placebo: 10.1 days; η^2^_p_= 0.130, power=0.198, respectively).

#### COVID-19 Biomarkers

Among biomarkers associated with COVID-19 risk, CRP exhibited a medium and significant difference between FoTv and Placebo groups across Days (η^2^_p_= 0.081, power=0.513; Table 3, Supplemental Table 2). There was a very large and significant decline in CRP in the FoTv group (η^2^_p_= 0.319, power=0.908; F = 11.735, p = 0.002), but only a large and non-significant decline in the Placebo group (η^2^_p_= 0.153, power=0.479; F_(1,23)_ = 3.831, p = 0.063). Similar Treatment Group by Day effect sizes were found for hs-cTnT and Ferritin (see Table 3 and Supplemental Table 2). There was a large and significant increase across days for hs-cTnT in the FoTv group (η^2^_p_= 0.188, power=0.619; F_(1,28)_ = 7.216, p = 0.012), but there was a small and non-significant decrease in the Placebo group (η^2^_p_= 0.041, power=0.154; F_(1,31)_ = 0.508, p = 0.482). Ferritin levels exhibited a large and significant decrease across days for the Placebo group (η^2^_p_= 0.268, power=0.774; F = 8.750, p = 0.007), but only a small and non-significant decrease in the FoTv group (η^2^_p_= 0.000, power=0.051; F = 0.000, p = 0.993).

#### SARS-CoV-2 Viral Load

There were no statistically significant differences between FoTv and Placebo groups across days for SARS-CoV-2 viral load (Days 0, 7, and 14) (η^2^_p_= 0.101, power=0.543; F_(2,83)_ = 2.609, p = 0.080). SARS-CoV-2 viral loads consistently declined across the study period (Day Effect: F_(2,84)_ = 169.739, p < 0.001) and there were no significant differences between the groups (Group main effect, F_(1,42)_ = 0.888, p=0.351; Table 3, Figure 3B). There were also no significant differences between the groups at Day 0, 7, or 14 (η^2^_p_= 0.001, 0.066, and 0.001, respectively; Figure 3B).

#### In Vitro Antiviral Assessment

We observed that Vero-TMPRSS2 cells bathed in media containing FoTv in DMSO (N=3, 91002±7451 relative light units; RLU) exhibited significantly reduced relative luminescence units (F_(1,4)_ = 22.395, p = 0.009), compared to cells bathed in media containing the same concentration of DMSO, as a negative control (N=3, 161933±24868 RLU), suggesting that FoTv reduced viral entry into the cells (see Figure 3C).

## DISCUSSION

This randomized, placebo-controlled Phase I clinical trial demonstrates that FoTv is safe, well tolerated, and feasible for the treatment of mild-to-moderate COVID-19 and provides evidence supporting further clinical investigation of mycelium-based interventions for viral disease. Aside from modest between-group mean differences in serum levels of adjusted eGFR (renal function), FoTv and Placebo serum levels of hepatic function, renal function, and coagulation were equally likely to transition from normal to abnormal levels across the treatment period (Table 2). Despite a high daily dosage (8 capsules, 3 times daily), compliance was remarkably high, with 99% of subjects completing the 14-day regimen and 96% reporting full adherence, suggesting treatment tolerability. This suggestion is supported by the safety data, with no sentinel evidence of cytokine storm in the FoTv subgroup, and no significant adverse events reported or meaningful differences detected in hepatic, renal, or coagulation biomarkers in the FoTv versus Placebo group. Given that the overall symptom burden was lower and less severe in the FoTv group, it suggests that any potential treatment-related side effects did not add to or exacerbate COVID-19 symptoms.

Although adjusted eGFR significantly decreased in the FoTv group from Day 0 to Day 14, mean values for both groups at both time points were well within the normal range for renal function (> 60 mL/min; Supplemental Table 1), with Day 14 mean adjusted eGFRs of 104.28 and 103.87 for FoTv and Placebo groups, respectively (Table 2).

Across the treatment period, participants in the FoTv group were no more likely to transition from normal to abnormal eGFR than were participants in the Placebo group (see overlapping confidence intervals in Table 2). Finally, concordant findings of FoTv’s safety were reported in a study of healthy individuals who took FoTv over a 4-day treatment period as an adjunct to COVID-19 vaccination^33^. Nevertheless, for participants with an active infection, caution should be employed when using FoTv at this dosage for a timeframe longer than 14 days. Future studies with longer treatment periods (and possibly lower dosage regimens) and in other patient populations and disease settings may be warranted.

Another COVID-specific safety consideration for FoTv relates to observations in the early days of the COVID-19 pandemic, that SARS-CoV-2 infection could suddenly and inexplicably trigger cytokine storm (signal event). Cytokine storm is a severe and potentially life-threatening immune overreaction to infection. Specifically, because SARS-CoV-2 initially suppresses the body’s early alarm system (i.e., type-1 interferon), viral reproduction proceeds unchecked by the immune system. It is only after viral loads reach a massive peak that the body then initiates an unchecked explosive immune response. Though intended to clear the virus, it instead induces a hyper-inflammatory state, marked by a failure to switch from innate to adaptive immunity, generation of a pro-inflammatory loop of cytokine release, tissue damage, and onset of potentially life-threatening symptoms^45^. Given that the risk for cytokine storm may be higher in those with a seemingly more robust (but arguably hyperactive or dysregulated) immune response, the known immune-enhancing effects of fungal polysaccharides generated speculation that therapeutic use of fungi might inadvertently trigger this event^44^.

However, we found no evidence of cytokine storm in the FoTv group. First, there were no adverse events reported (for any participant). Second, the number and severity of symptoms declined faster over the treatment period in the FoTv than the Placebo group (Table 3, Figure 3A). Finally, because CRP is the primary hepatic biomarker triggered by IL-6 during cytokine storm, the large, significant decline in CRP in the FoTv group directly contradicts the presence of systemic cytokine storm (Table 3). Moreover, a clinical trial of COVID-19 patients found a fungal-derived treatment could actually decrease inflammatory markers IL-6 and D-dimer^46^. This idea is reinforced by the fact that polypore fungi contain various bioactive peptides^47^ which can regulate cytokine responses, enhancing the shift from innate to adaptive immunity, thereby optimizing the overall immune response^48^.

It is notable that several COVID-19 biomarkers significantly changed across the treatment period in a manner that was different for the FoTv versus Placebo groups (Table 3). Specifically, for CRP there was a larger decline in FoTv than in Placebo, whereas for hs-cTnT there was an increase for FoTv and no change for Placebo, and for Ferritin, no change for FoTv and a decrease for Placebo. While biomarker trajectories differed between the groups, clinical symptoms declined in both groups. However, decreases in symptom number and severity were more pronounced in the FoTv group (Figure 3). These seemingly paradoxical findings may suggest that FoTv, rather than solely and broadly suppressing inflammatory pathways, functions in a more specific and nuanced immunomodulatory manner ^49^.

These three biomarkers reflect very different pathways, which could possibly explain the biomarker and clinical findings. CRP, mediated by IL-6 and TNF-alpha, more purely reflects systemic inflammation, and specifically in acute COVID-19^50^. Given the greater reduction in clinical symptoms in the FoTv group than in Placebo, the greater reduction in CRP in the FoTv group may indeed reflect an anti-inflammatory effect of fungi^46^ and specifically Fo ^21,51^. Ferritin, while considered an acute phase reactant, also reflects sequestration of iron to protect the host from virally induced cellular stress in the face of macrophage activation. This suggests that stable Ferritin levels in the FoTv group (vs. a reduction in Placebo), could reflect FoTv’s ability to protect tissue from iron-induced oxidative stress during macrophage-mediated viral clearance^52^. For the modest increase in hs-cTnT in the FoTv group (vs. no change in Placebo), this elevation may cause concern for myocardial strain. However, the absence of any adverse events and the faster resolution of clinical symptoms, particularly muscle aches, argues against cardiotoxicity and in favor a non-cardiac reason for the increase in hs-cTnT. Further, we employed the high-resolution hs-cTnT assay, which can detect not only myocardial but also regenerating skeletal muscle fibers^53–55^. However, because cardiac-specific Troponin-I and skeletal muscle-specific enzymes were not measured, future studies are needed to confirm whether the increased hs-cTnT reflected subtle, enhanced skeletal muscle tissue repair (consistent with faster reduction in muscle aches in the FoTv than in the Placebo group) or, less likely, a subclinical cardiac muscle damage (inconsistent with the overall pattern of improvement in other COVID biomarkers and symptoms in the FoTv group).

The finding that the FoTv group exhibited fewer (and less severe) COVID-19 symptoms across the study period versus Placebo (Figure 3A, Table 3), and that the effect sizes were moderately strong, merits additional discussion. The largest effect sizes were for symptoms most significantly reduced in severity were sore throat, cough, and runny nose (which all have an inflammatory component). Among symptoms of COVID-19, FoTv’s effect on cough was one of the largest. This is notable because cough is the primary driver of person-to-person viral transmission, due to its unique ability to forcefully expel viral particles in high concentrations, with particles travelling further and remaining in the air longer (vs. breathing or talking)^56,57^.

Although the study was not designed to examine symptom effects according to vaccination status, we observed that both vaccinated and unvaccinated participants who received FoTv (vs. Placebo) exhibited a reduction in symptoms, with a strong trend suggesting a more robust effect of FoTv in the unvaccinated group. Specifically, the unvaccinated FoTv group (vs. unvaccinated Placebo) appeared to “turn the corner faster”, i.e., 5.1 days faster (large effect size). In contrast, the vaccinated FoTv group (vs. vaccinated Placebo) experienced a 50% reduction in symptoms only 1.3 days faster for FoTv than Placebo (small effect size). These findings may have public health significance as they suggest FoTv might serve both as a suitable treatment for viral infection and method for reducing person-to-person transmission, at a time when vaccines may not yet be broadly available or may not be accepted by some individuals.

Given that FoTv also appears to reduce vaccine-related side effects when used as an adjunct to COVID-19 vaccines (Pfizer, Moderna, Johnson & Johnson)^33^, our findings suggest that FoTv might have the ability to reduce symptoms of immune response to active viral infection as well as induced viral challenge. Preclinical findings suggest that *Phellinus linteus*, a polypore fungus in the same class as FoTv (i.e., Agaricomycetes), enhanced vaccination activity against H5N1^35^. If these effects are not specific to SARS-CoV-2 and COVID-19 vaccination and were to be applied generally to other infectious diseases and vaccinations, FoTv might potentially have even broader public health impact. Given the increased concerns about the H5N1 bird flu contagion, more research is warranted to explore these possible benefits. We are currently expanding the scope of our preclinical investigations to assess FoTv against high-consequence viral threats—including Nipah, Ebola, avian and mammalian influenza, and hantaviruses—in BSL-3 and BSL-4 Select Agent laboratories.

In contrast with the differences in symptoms between the FoTv and Placebo groups, these groups did not exhibit any differences over time in SARS-CoV-2 viral RNA levels according to nasal swab PCR. However, because PCR only measures total viral RNA, it cannot distinguish infectious from noninfectious viral material, making it difficult to determine whether FoTv may have reduced viral infectivity without substantially altering RNA levels. This distinction is particularly relevant because the FoTv group showed reduced clinical burden (Figure 3A) despite no apparent difference in nasal swab viral RNA levels (Figure 3B). It is also consistent with prior reports that both Fo and Tv exhibit antiviral activity against other live viruses, including influenza A subtypes H5N1 and H3N2^8,15,16,23,24^. We therefore tested FoTv in an *in vitro* assay, where it significantly reduced infection by SARS-CoV-2 pseudovirus in cells (Figure 3C). In contrast to the similar viral RNA measurements observed in nasal swabs for FoTv and Placebo groups, the *in vitro* results support antiviral activity of FoTv and raise the possibility that some of its clinical effects may have involved reduction of infectious virus not detected by PCR testing alone. For example, reduced cellular infection, by reducing viral biomass and tissue damage, is known to result in lower downstream release of proinflammatory cytokines ^58–60^. Future clinical trials incorporating plaque assays or other infectivity-based methods will likely be needed to determine whether FoTv reduces SARS-CoV-2 or other viral infection in humans.

There are numerous reasons why fungal mycelial products like FoTv may be practical for widespread, standardized use as adjuncts to standard care COVID-19 and other viral diseases. Fungi [1] are chemically complex and biosynthesize numerous compounds and may work synergistically to address several elements of viral infection through both innate and adaptive immune responses; [2] may constitute a safe, natural, sustainable, scalable, low-cost, and feasible treatment to mitigate symptoms; and [3] allow for rapid and high-volume production because the technology employs existing infrastructure for aseptic cultivation using solid-state fermentation. Population-scale medical use of mushroom fruit bodies is likely infeasible due to impracticality of controlled cultivation, potential for contamination with insects and microorganisms^61^, and concerns about overharvesting, particularly of agarikon, which has been listed as Endangered by the International Union for Conservation of Nature^62^. In contrast, industrial production of mycelia from fungi, such as Fo and Tv, is a realizable, standardizable, and scalable strategy for population-level treatment of viral diseases with pandemic potential. Finally, although not as strictly regulated as drugs, which may only receive FDA approval following demonstrations of safety and efficacy in clinical trials, over-the-counter Fo and Tv have been broadly consumed across the United States for over 10 years. Consistent with standard production practices at Fungi Perfecti, LLC, which provided the treatments used in this clinical trial, both FoTv and Placebo were subjected to rigorous quality assurance testing to ensure product fidelity, identity of the cultivated species, and to confirm the absence of microbiological contamination (see Supplemental Table 3).

This study had several limitations warranting discussion. First, while we selected a particular dosage and duration, alternate regimens might have resulted in different effects on symptoms and other outcomes. Second, the rapidly changing nature of the COVID-19 pandemic and administrative delays that followed delayed the start of the study to a time that was less advantageous for identifying FoTv’s effect on morbidity from COVID-19. Specifically, the protocol was designed in March 2020, when COVID-19 hospitalization rates up to 26% were reported^63^; however, recruitment only began after the winter 2020/2021 COVID-19 peak and initial vaccine deployment, preventing in-depth examination of the effects of FoTv on COVID-19 illness itself, including important outcomes such as hospitalization rate and ER visits. Had the study been launched earlier, more unvaccinated patients infected with the higher-morbidity SARS-CoV-2 Alpha variant may have been recruited, and thus FoTv treatment might have demonstrated even greater reductions in symptom count and severity, as well as ER visits and hospitalizations. Finally, even though we employed *in vitro* methods that are standard in the field, we tested FoTv on cells infected with a SARS-CoV-2/vesicular stomatitis virus (VSV) pseudotyped virus (rather than SARS-CoV-2 itself) and this test was conducted on non-human mammalian cells. And, as is the case with all cell culture research, our *in vitro* findings may not generalize to humans.

## Conclusions

In this double-blind, placebo-controlled, randomized clinical trial conducted under FDA oversight, treatment with a combination of Agarikon and Turkey tail mycelia, was shown to be safe, well-tolerated, and feasible in individuals with mild-to-moderate acute infection with SARS-CoV-2 who were self-quarantining at home. Exploratory analyses indicated large effects on COVID-19 symptoms (reducing both their number and severity), particularly cough, runny nose, and sore throat (which, in theory, could reduce disease transmission), muscle aches, and other symptomatology. These findings were concordant with the significant reduction in short-term COVID vaccination side effects^33^ observed in individuals who had not been previously infected with or vaccinated against SARS-CoV-2. Together with the COVID-19 serum biomarker results (which suggest immunomodulatory effects across several markers), our findings suggest that FoTv modulation of the immune response to active viral infection and induced viral challenge may result from common, context-dependent innate immune-mediated mechanisms involving pathways beyond β-glucan-associated Dectin-1, including TLRs, NF-κB signaling, and cytokine activity, particularly the IL-1 axis, as demonstrated preclinically for agarikon mycelium^31^. This suggestion is further supported by the results of the *in vitro* study that found that pre-infection exposure to FoTv reduced SARS-CoV-2 cellular infection. Taken together, these studies raise the intriguing possibility that, in the event of a future viral pandemic, FoTv might: (a) provide an effective means for reducing symptom severity in the absence of any proven pharmaceutical or other treatments, (b) help break the chain of transmission by reducing both cellular infection and the spread of viral particles through coughing, and (c) when vaccines become available, reducing vaccine hesitancy and possibly increasing the magnitude and duration of antibody response. If so, these studies could elucidate a strategy for a novel two-phase public health pandemic intervention for disease with pandemic potential: use of a botanical immunomodulatory product both during the early critical months of an outbreak to ameliorate symptoms and reduce spread and, later, after vaccines become available, to increase their uptake and effectiveness.

The same degree of symptom reduction was observed regardless of vaccination status (perhaps with a greater and more rapid reduction in unvaccinated participants). Moreover, polypore or polypore-like fungi also appear to be effective adjuncts to COVID-19 vaccination in humans^33^ and influenza A/H5N1 vaccination in mice^35^. Therefore, FoTv’s potential as a therapeutic for active COVID-19 infection adds to these prior vaccination findings and raises the intriguing possibility that FoTv or other polypore (or polypore-like) fungi could also serve as adjuncts to vaccines for viruses beyond SARS-CoV-2. These could potentially include influenza A viruses such as human H3N2 and avian H5N1. Future studies of FoTv may explore different dosing regimens and durations in the treatment of COVID-19 or other viral diseases with pandemic potential where vaccinations may not be available or acceptable by significant portions of the population.

## Data Availability

All data produced in the present study are available upon request from the corresponding author.

## ARTICLE INFORMATION

### Conflict of Interest Disclosures: None

#### Disclaimer

None.

#### Data Sharing Statement

Data are available upon request from the corresponding author.

#### Authors’ Contributions

GS, AS, SG, SW, and HAC contributed to conceptualization; SG, TS, PS, NG, and DB contributed to data curation; SG and CNS carried out the formal analysis; GS and LM contributed to funding acquisition; TS, PS, and SG contributed to investigation; GS, SG, AS, LK, DB, and HAC contributed to methodology; TS, AS, PS, LM, and GS contributed project administration; TS, SG, PS, and RD contributed resources; SG contributed software; GS, TS, AS, LM, and DS supervised the work; SG and CNS validated the data; CNS, GS, and DB contributed to visualization; GS, CNS, AS, ZB, CB, DB, and HAC wrote the original draft; and all authors contributed to reviewing and editing the final draft.

## Acknowledgements

We are thankful for the visionary and pioneering work of Paul Stamets, whose contributions were critical to the development of the MACH-19 studies. We would also like to acknowledge Lee Stein, Esq., whose advocacy and encouragement made the MACH-19 studies possible.

## Ethics approval and consent to participate

Written informed consent was obtained for all patients included in this clinical trial. This study was conducted in accordance with the ethical guidelines of the Declaration of Helsinki and approved by the UCSD Institutional Review Board: 200633-1a.

## Consent for publication

Not applicable.

