## Supplemental Information for "Safety, Feasibility, and Preliminary Clinical Findings of an Oral Polypore Fungi Combination with Mild-to-Moderate COVID-19: A Randomized, Placebo-Controlled Phase I Clinical Trial"

| Supplemental Table 1. Cut off scores for identifying normal and abnormal values for liver function, renal function, and coagulation | | | | | | | |
| --- | --- | --- | --- | --- | --- | --- | --- |
|  | UCSD | |  | UCLA | |  |  |
|  | Normal | Abnormal |  | Normal | Abnormal |  | Unit |
| *Liver Function* |  |  |  |  |  |  |  |
| Total Protein | 6.0 - 8.0 | < 6 or > 8 |  | 6.1 - 8.2 | < 6.1 or >8.2 |  | g/dL |
| Albumin | 3.5 - 5.2 | < 3.5 or > 5.2 |  | 3.9 - 5.0 | < 3.9 or > 5.0 |  | g/dL |
| Alkaline Phosphatase (ALP) | ≤ 129 | > 129 |  | ≤ 113 | > 113 |  | U/L |
| Aspartate Aminotransferase (AST) | ≤ 40 | > 40 |  | ≤ 62 | > 62 |  | U/L |
| Alanine Transaminase (ALT) | ≤ 41 | > 41 |  | ≤ 70 | > 70 |  | U/L |
| Total Bilirubin | <1.2 | ≥ 1.2 |  | 0.1 - 1.2 | < 0.1 or > 1.2 |  | mg/dL |
| *Renal Function* |  |  |  |  |  |  |  |
| Adjusted glomerular filtration rate  (Adj. eGFR) | > 60 | ≤ 60 |  | > 60 | ≤ 60 |  | mL/min |
| Sodium | 136 - 145 | < 136 or > 145 |  | 135 - 146 | < 135 or > 146 |  | mmol/L |
| Chloride | 98 - 107 | < 98 or > 107 |  | 96 - 106 | < 96 or > 106 |  | mmol/L |
| Blood Urea Nitrogen (BUN) | 8 - 23 | < 8 or > 23 |  | 7 - 22 | < 7 or > 22 |  | mg/dL |
| *Coagulation* |  |  |  |  |  |  |  |
| Prothrombin Time | 9.7 – 12.5 | < 9.7 or > 12.5 |  | 9.7 – 12.5 | < 9.7 or > 12.5 |  | sec |
| Activated partial thromboplastin time (APTT) | 27 - 36 | < 27 or > 36 |  | 27 - 36 | < 27 or > 36 |  | sec |
| International Normalized Ratio (INR) | Not provided by laboratory | |  | Not provided by laboratory | |  | N/A |
| *Note*. Adj. eGFR = 142 X minimum (standardized serum creatinine/K OR 1)^α^ X maximum(standardized serum creatinine/K OR 1)^-1.200^ X 0.9938^Age^ X 1.012 [if female]. Serum creatinine = mg/dL. K = 0.7 (females) or 0.9 (males). α = -0.241 (females) or -0.302 (males). Equation based on National Kidney Foundation guidelines (https://www.kidney.org/ckd-epi-creatinine-equation-2021-0). | | | | | | | |

| Supplemental Table 2. Mixed Effects Models Examining Treatment Groups across Days for Efficacy Outcomes | | | | | | | | |
| --- | --- | --- | --- | --- | --- | --- | --- | --- |
|  |  | Treatment Group  Main Effect | | |  | Treatment Group  by Day Interaction | | |
| COVID-19 Symptoms |  | F value | p value | df |  | F value | p value | df |
| Sore throat |  | 3.349 | 0.072 | 1,66 |  | 46.928 | <0.001 | 1,697 |
| Cough |  | 2.156 | 0.148 | 1,54 |  | 18.931 | <0.001 | 1,697 |
| Muscle aches |  | 2.139 | 0.149 | 1,54 |  | 10.826 | 0.001 | 1,697 |
| Headache |  | 6.017 | 0.017 | 1,56 |  | 8.932 | 0.003 | 1,697 |
| Runny nose |  | 5.174 | 0.027 | 1,58 |  | 5.264 | 0.022 | 1,697 |
| Stuffy nose |  | 0.028 | 0.869 | 1,55 |  | 4.324 | 0.038 | 1,697 |
| Fatigue |  | 0.144 | 0.706 | 1,52 |  | 2.990 | 0.084 | 1,697 |
| Loss of taste |  | 0.702 | 0.406 | 1,51 |  | 2.024 | 0.155 | 1,697 |
| Fever |  | 1.742 | 0.191 | 1,76 |  | 1.909 | 0.168 | 1,697 |
| Loss of smell |  | 0.035 | 0.852 | 1,50 |  | 1.728 | 0.189 | 1,697 |
| Shortness of breath upon exertion |  | 0.004 | 0.951 | 1,47 |  | 0.290 | 0.591 | 1,613 |
| Shortness of breath |  | 3.647 | 0.061 | 1,55 |  | 0.010 | 0.919 | 1,697 |
| Number of Symptoms |  | 4.662 | 0.035 | 1,53 |  | 7.835 | 0.005 | 1,697 |
| Severity of Symptoms |  | 3.003 | 0.089 | 1,52 |  | 16.560 | <0.001 | 1,697 |
| COVID-19 Biomarkers |  |  |  |  |  |  |  |  |
| hs-cTnT^1^ |  | 4.671 | 0.033 | 1,114 |  | 5.016 | 0.029 | 1,62 |
| Ferritin^2^ |  | 0.505 | 0.479 | 1,99 |  | 4.686 | 0.035 | 1,49 |
| CRP |  | 4.027 | 0.048 | 1,79 |  | 4.280 | 0.044 | 1,48 |
| D-dimer^2^ |  | 2.001 | 0.160 | 1,132 |  | 1.175 | 0.283 | 1,62 |
| ESR |  | 0.594 | 0.444 | 1,69 |  | 0.090 | 0.765 | 1,41 |
| LDH |  | 0.853 | 0.358 | 1,78 |  | 0.039 | 0.884 | 1,48 |
| SARS-CoV-2 Viral Load3 |  | 0.888 | 0.351 | 1,42 |  | 2.609 | 0.080 | 2,83 |
| Note. ^1^Data available only from UCSD site. ^2^Recruitment site laboratories used different standards, therefore z scores were used. CRP, C-reactive protein; ESR, estimate sedimentation rate; LDH, lactate dehydrogenase; hs-cTnT, high-sensitivity Troponin-T Gen 5. | | | | | | | | |

Supplemental Figure 1. Symptom Count and Symptom Severities plotted separately for Vaccinated and Unvaccinated participants.

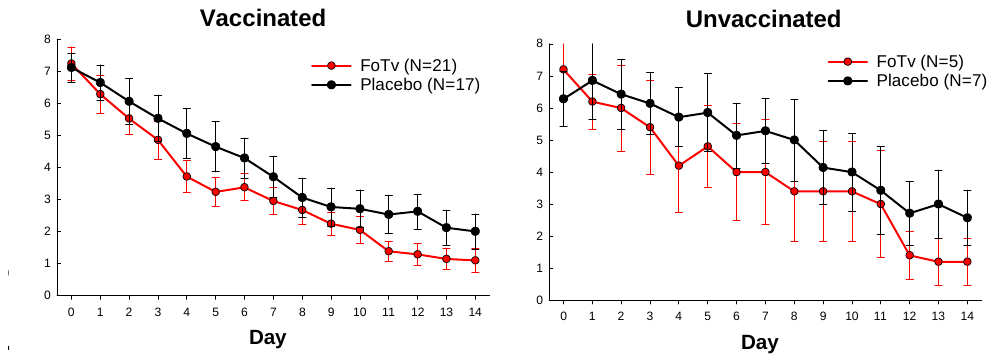

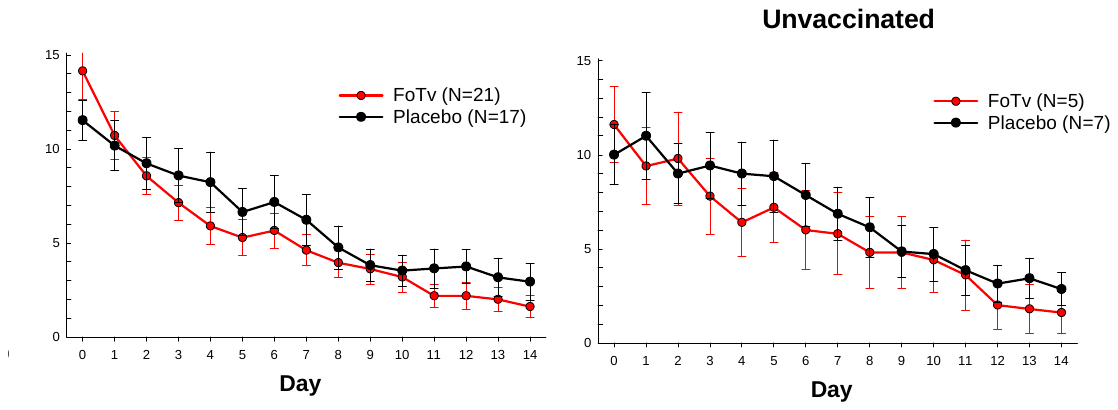

| Supplemental Table 3. FoTv Capsule Microbiological Testing | | |
| --- | --- | --- |
| Microbe | Method | Result |
| Salmonella | USP | Absent/10 gram |
| Listeria | LISTBAX | Not Detected/25 g |
| Aerobic Plate Count | AOAC 990.12 | < 10 cfu/gram |
| Yeast & Mold | AOAC 997.02 | < 10 cfu/gram |
| Coliforms | AOAC 991.14 | < 10 cfu/gram |
| E. coli | AOAC 991.14 | Not Detected |
| Enterobacteriaceae | Petrifilm | < 10 cfu/gram |
| *Note.* Microbiological testing results conducted by the capsule co-manufacturers had the best possible outcomes. cfu/gram, colony-forming unit per gram. | | |
